# Prevalence and Longevity of SARS-CoV-2 Antibodies in Healthcare Workers: A Single Center Study

**DOI:** 10.1101/2020.10.09.20210229

**Authors:** Michael Brant-Zawadzki, Deborah Fridman, Philip A. Robinson, Matthew Zahn, Clayton Chau, Randy German, Marcus Breit, Elmira Burke, Jason R. Bock, Junko Hara

**Affiliations:** Hoag Center for Research and Education, Hoag Memorial Hospital Presbyterian, Newport Beach, California, United States; Infection Prevention, Hoag Memorial Hospital Presbyterian, Newport Beach, California, United States; Orange County Health Care Agency, Santa Ana, California, United States; Laboratory Administrative Services, Hoag Memorial Hospital Presbyterian, Newport Beach, California, United States; Hoag Family Cancer Institute, Hoag Memorial Hospital Presbyterian, Newport Beach, California, United States; Quality Management, Hoag Memorial Hospital Presbyterian, Newport Beach, California, United States; Medical Care Corporation, Newport Beach, California, United States

## Abstract

Understanding SARS-CoV-2 antibody prevalence as a marker of prior infection in a spectrum of healthcare workers (**HCWs**) may guide risk stratification and enactment of better health policies and procedures.

The present study reported on cross-sectional study to determine the prevalence and longevity of SARS-CoV-2 antibodies in HCWs at a regional hospital system in Orange County, California, between May and August, 2020.

Data from HCWs (n=3,458) were included in the analysis. Data from first responders (n=226) were also analyzed for comparison. A blood sample was collected at study enrollment and 8-week follow-up. Information on job duties, location, COVID-19 symptoms, polymerase chain reaction test history, travel since January 2020, and household contacts with COVID-19 was collected. Comparisons to estimated community prevalence were also evaluated.

Observed antibody prevalence was 0.93% and 2.58% at initial and 8-week follow-up, respectively, for HCWs, and 5.31% and 4.35% for first responders. For HCWs, significant differences (*p* < .05) between negative vs. positive at initial assessment were found for age, race, fever, and loss of smell, and at 8-week follow-up for age, race, and all symptoms. Antibody positivity persisted at least 8 weeks in this cohort. Among 75 HCWs with self-reported prior PCR-confirmed COVID-19, 35 (46.7%) were antibody negative. Significant differences between negative vs. positive were observed in age and frequency of symptoms.

This study found considerably lower SARS-CoV-2 antibody prevalence among HCWs compared with prior published studies. This may be explained by better safety measures in the workplace, heightened awareness inside and outside of the workplace, possibly lower susceptibility due to innate immunity and other biological heterogeneity, and low COVID-19 prevalence in the community itself. HCWs with initial positive results had persistent positive serologies at 8 weeks. Further research is warranted to investigate factors influencing such lower prevalence in our HCWs.

## INTRODUCTION

Since first reported in Wuhan, China in December 2019, coronavirus disease 2019 (**COVID-19**) has given rise not only to a tremendous healthcare and socioeconomic crisis worldwide, but also to unprecedented psychological trauma to the world community, including healthcare workers (**HCWs**) and individuals, with a wide-ranging downstream impact and an anticipated prolonged recovery.

COVID-19’s extraordinary infectivity, given its novel nature and pre-symptomatic transmission, has fueled its wide and wild spread across and within countries, with confirmed cumulative cases of 7.6 million in United States and 36.3 million worldwide as of October 8, 2020. A recent review article reports that approximately 40-45% of those infected with SARS-CoV-2 could be asymptomatic for an extended period of time (e.g., beyond 14 days) or never develop symptoms, suggesting a much wider spread of the virus than confirmed cases indicate [1]. It is estimated that a ten-fold presence of infection exists for every confirmed case [2-4]. Some sub-sampled confined cohorts demonstrated asymptomatic prevalence as high as 96% [1,5]. Early anecdotal media reports suggested that HCWs were particularly vulnerable to infection. While great effort has been put into development of SARS-CoV-2 vaccines, it is of great importance to better understand the extent of transmission within health facilities, and the susceptibility of the health-care workforce to infection, so that better and prioritized preventive strategies can be developed and deployed.

Sero-surveillance studies have been conducted to estimate SARS-CoV-2 antibody prevalence in various countries and settings, including sero-surveillance among blood donors [6-8]. Such estimates help better understand the nature of total numbers of infected individuals to estimate the true infection mortality rate (vs. the case fatality rate). Equally if not more important, true infection prevalence and its change over time would better explain the nature of asymptomatic and pre- or peri-symptomatic transmissions, environmental differences, and possibly duration of antibody presence. This is particularly of interest in acute health care settings.

However, previously reported results of sero-surveillance have varied greatly due to factors including sample size and geography (e.g., high active infection zones vs. low), ranging from 57% prevalence in Bergamo, Italy [9], 12.5% in New York State [10], down to 4.7 % in Los Angeles County [11] and 2.8% in Santa Clara County [12], California. The reliability of some of the early methodology for measuring antibodies might have also contributed to these varying results [13].

For HCWs, SARS-CoV-2 antibody prevalence has been sparsely reported but also with similar limitations as to sample size, wide range of results, timing of sampling, and variable methodologies. Such prevalence across different hospitals or healthcare networks ranges from 89.3% in Wuhan, China (n = 424) [14], 35.8% in New York City (n = 285) [15], 13.7% in the greater New York city area (n=40,329) [16], to 7.4% in Milan, Italy [17], and 2.67% in Denmark (n = 28,792) showing higher association between positivity and job duties, younger age (<30), and self-reported suspicion of prior COVID-19 exposure and prior positive PCR testing [18]. Determining such prevalence in a wide spectrum of HCWs, using a validated and accurate serum assay and repeated sampling over time to measure duration of antibody presence, may help stratify the work force for risk, limit transmission across different healthcare settings, enact better mitigation processes and procedures, and possibly better prioritize future vaccine delivery to front line workers.

The present study reports and expands [19] on the sero-surveillance conducted among HCWs at Hoag Memorial Hospital Presbyterian, Orange County, California, and subsequent follow-up at 8 weeks. An additional smaller sample of sero-surveillance among first responders (e.g., fire captains, police officers) in Orange County, CA, as well as antibody positive prevalence calculated from community physician orders, are also reported for comparative purposes. The study was conducted between May and Aug, 2020.

## METHODS

### Subject Recruitment

Institutional review board (**IRB**) approval was obtained for this study from Providence St. Joseph Health IRB (IRB# 2020000337). Study HCW subjects were recruited by direct email notifications to the entire employee workforce (6,500+ individuals) and independent medical staff (1,600+ physicians), whose work locations include two hospital campuses, nine health centers, thirteen urgent care locations, and other clinical and administrative facilities all within approximately a 20-mile radius. Similarly, study subjects from first responders were recruited from fire and police departments in Orange County, CA, by direct email notifications.

### Enrollment and Data Collection

Informed consents were obtained in person originally, then transitioned to electronic consent format starting June 19, 2020. Those who were enrolled through in-person consent were surveyed for job duties, location, COVID-19 symptoms, a self-reported polymerase chain reaction (**PCR**) test history with test date if available, travel record since January 2020, and existence of household contacts with COVID-19. Those who were enrolled through electronic consent format answered the same survey online at the time of consenting. The COVID-19 symptoms survey included fever, sore throat, cough, runny nose, and loss of sense of smell, with loss of taste added at 8-week follow-up. Using reported job duties and locations, each HCW subject was classified into a) high (e.g., MD, RN, PA, emergency care tech, ICU tech), b) medium (e.g., therapist, phlebotomist, medical tech), or c) low (e.g., admin, coder, billing, lab tech/scientist, IT) risk groups to approximate levels of direct exposure to COVID-19 patients.

### Blood Sample Collection

The first blood sample (∼5ml) was collected for serum analysis for IgG antibodies to SARS-CoV-2 at the time of in-person consent, or following electronic consent at two main hospital campuses. With the exception of 16 subjects, blood sample collection was within 7 days of electronic consent (*M* = 1.77, *SD* = 1.83). Eight weeks after the first blood sample, the second sample was collected.

### IgG Antibodies to SARS-CoV-2 Analysis

Serum analysis for IgG antibodies to SARS-CoV-2 utilized the Ortho Clinical Diagnostics VITROS® XT 7600 platform. A 5 ml peripheral draw venous blood sample was collected from each subject into a gold top serum separator vacutainer tube (BD Medical). Samples were centrifuged within 2 hours of collection at 4500 RPM for 5 minutes (RCF 3060). Aliquots were analyzed with calibrated lots of Anti-SARS-CoV-2 IgG Reagent Pack on the VITROS® XT 7600 according to manufacturer’s instructions for use [20]. Positive and negative quality controls were run daily prior to sample analysis (Ortho Diagnostics Anti-SARS-CoV-2 IgG Control). At the time of writing, this IgG test is approved only for use under the Food and Drug Administration’s Emergency Use Authorization (EUA), and is also used in CDC studies [21].

Manufacture sensitivity and specificity claims for the Ortho Clinical Diagnostics VITROS Anti-SARS-CoV-2 IgG assay is 100% (407/407) negative agreement (95% CI: 99.1–100.0%) in 407 presumed SARS-CoV-2 antibody negative subjects and 87.5% (42/48) positive agreement (95% CI: 74.8–95.3%) in 48 PCR positive subjects with days from positive PCR ranging from 1 day to 22 days and days from onset of symptoms ranging from 12 to 32 days. In-house validation studies were conducted with 35 samples from subjects with a known positive SARS-CoV-2 PCR test a mean of 43 days out from positive PCR test date (range 38-48 days), and 50 samples from subjects with a known negative SARS-CoV-2 PCR test. Of 31 PCR samples, 29 were positive for SARS-CoV-2 IgG antibody. All 50 of the PCR negative samples were SARS-CoV-2 IgG antibody negative. Thus, sensitivity of 93.6% (95%CI: 78.6–99.2%) and specificity of 100% (95% CI: 92.9–100.0%) were calculated for the Ortho Diagnostics VITROS Anti-SARS-CoV-2 IgG assay in run our laboratory on the Ortho Clinical Diagnostics VITROS® XT 7600 automated instrument platform, and adopted in this study.

### Data Analysis

Data were examined for HCWs and first responders at first and second blood draw results, each comparing antibody negativity vs. positivity. Nonparametric tests for group differences were performed for demographics and five symptoms of COVID-19 at the first blood draw, with an additional one symptom at the second blood draw. The effect of occupational risk was also evaluated for HCWs. Mann-Whitney U tests were used for assessing group difference in age, and a series of one-sided Fisher’s exact tests were used for the remaining categorical factors; for group differences in race (a 7×2 table) and occupational risk (a 3×2 table), the Mehta-Patel algorithm was applied [22]. A value of p < .05 was used for statistical significance. For all analyses, the Stata statistical software package, edition 15, was used [23].

## RESULTS

After excluding subjects for missing symptoms data, the final analyses included 3,458 subjects from the first blood draw and the subset of those who returned for the second draw (n = 2,754; 79.6% return rate) for HCWs, and 226 subjects from first blood draw and the subset of those who returned for the second draw (n = 92; 40.7% return rate) for first responders.

Among HCWs’ initial blood draw, 32 antibody positive cases (3,426 negative) were identified, with an observed prevalence of 0.93% (exact binomial 95% CI = 0.63% - 1.30%). Accounting for test sensitivity (93.6%) and specificity (100%), an adjusted prevalence of 0.98% (exact binomial 95% CI = 0.68% - 1.37%) was calculated, indicating 34 positive cases (3,424 negative) after adjustment. At their 8-week follow-up blood draw (n=2,754), 71 antibody positive cases (2,683 negative) were identified, with an observed prevalence of 2.58% (exact binomial 95% CI= 2.02% - 3.24%). Of the original 32 positive subjects, 28 remained positive (4 did not return for the second blood draw) with additional 43 new cases during an 8-week period (**Table 1a**). An adjusted prevalence of 2.76% (exact binomial 95% CI = 2.18% - 3.44%) was calculated, indicating 76 positive cases (2,678 negative) after adjustment. **Table 2** summarizes HCW sample characteristics and group differences.

**Table 1.** Sample Size Summary.

(a) Healthcare Workers
| 1st Draw Antibody Results | N | 2nd Draw Antibody Results | N |
| --- | --- | --- | --- |
| Negative | 3,426 | Negative | 2,683 |
|  |  | Positive | 43 |
|  |  | Did Not Return | 700 |
| Positive | 32 | Negative | 0 |
|  |  | Positive | 28 |
|  |  | Did Not Return | 4 |
| Total | 3,458 | Total Returned | 2,754 |

(b) First Responders
| 1st Draw Antibody Results | n | 2nd Draw Antibody Results | n |
| --- | --- | --- | --- |
| Negative | 214 | Negative | 88* |
|  |  | Positive | 3 |
|  |  | Did Not Return | 124 |
| Positive | 12 | Negative | 0 |
|  |  | Positive | 1 |
|  |  | Did Not Return | 11 |
| Total | 226 | Total Returned | 92 |

\* One subject at 2nd Draw was missing at 1st Draw.

**Table 2.**
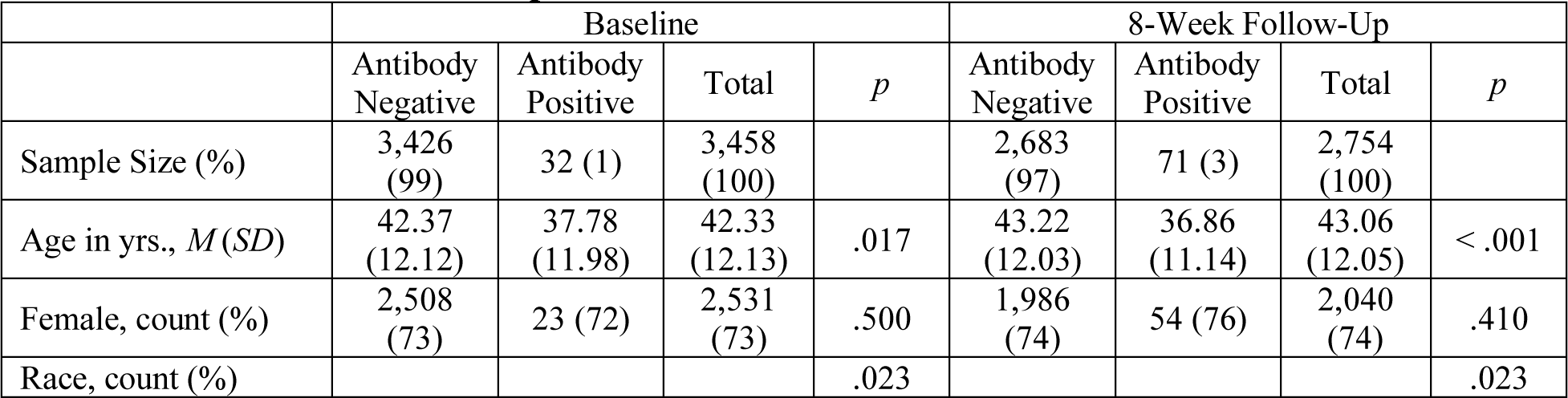
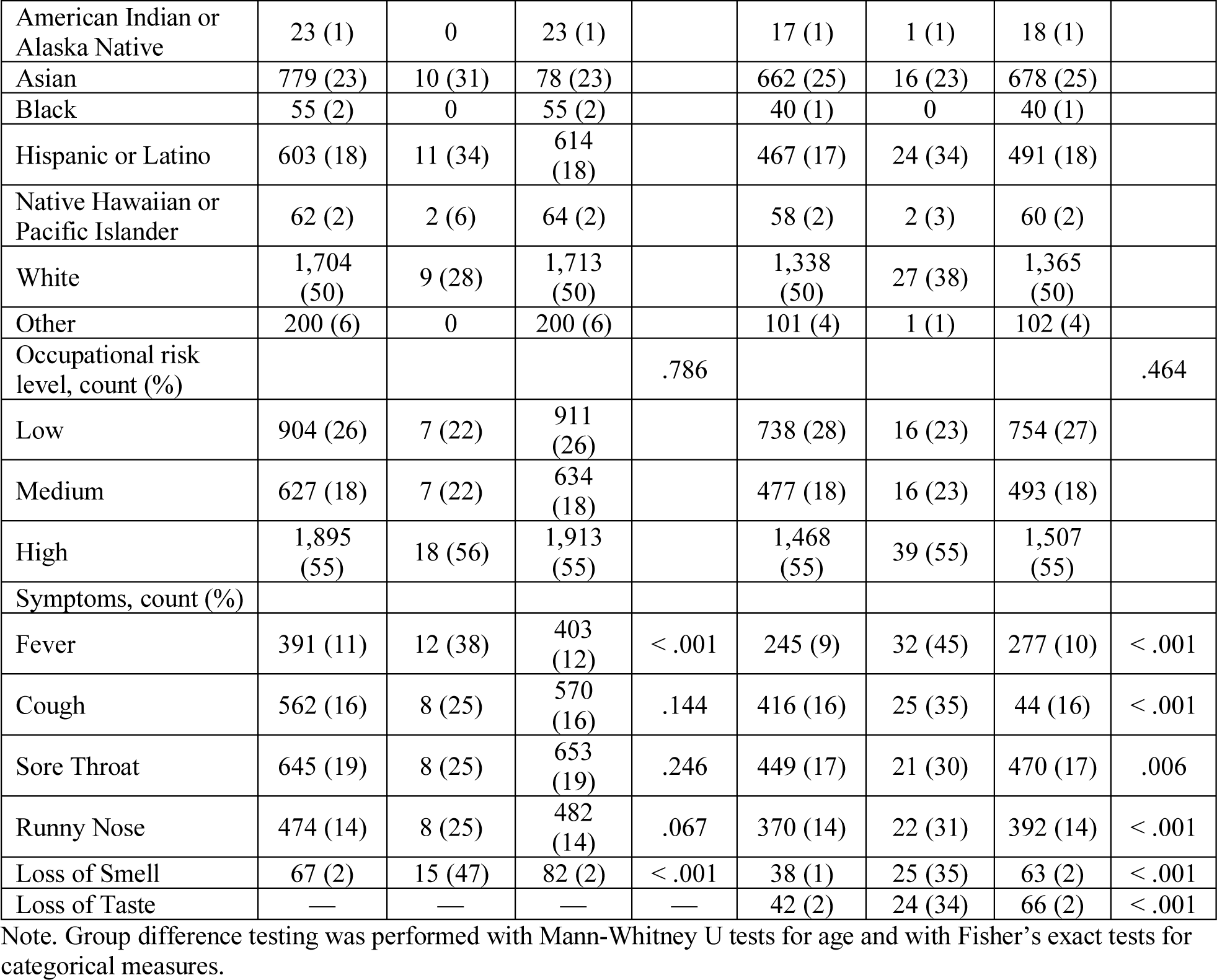
Sample Characteristics and Group Differences for Healthcare Workers at Baseline and 8-Week Follow-Up Assessments.

|  | Baseline |  |  |  | 8-Week Follow-Up |  |  |  |
| --- | --- | --- | --- | --- | --- | --- | --- | --- |
|  | Antibody Negative | Antibody Positive | Total | *p* | Antibody Negative | Antibody Positive | Total | *p* |
| Sample Size (%) | 3,426 (99) | 32 (1) | 3,458 (100) |  | 2,683 (97) | 71 (3) | 2,754 (100) |  |
| Age in yrs., *M* (*SD*) | 42.37 (12.12) | 37.78 (11.98) | 42.33 (12.13) | .017 | 43.22 (12.03) | 36.86 (11.14) | 43.06 (12.05) | < .001 |
| Female, count (%) | 2,508 (73) | 23 (72) | 2,531 (73) | .500 | 1,986 (74) | 54 (76) | 2,040 (74) | .410 |
| Race, count (%) |  |  |  | .023 |  |  |  | .023 |
| American Indian or Alaska Native | 23 (1) | 0 | 23 (1) |  | 17 (1) | 1 (1) | 18 (1) |  |
| Asian | 779 (23) | 10 (31) | 78 (23) |  | 662 (25) | 16 (23) | 678 (25) |  |
| Black | 55 (2) | 0 | 55 (2) |  | 40 (1) | 0 | 40 (1) |  |
| Hispanic or Latino | 603 (18) | 11 (34) | 614 (18) |  | 467 (17) | 24 (34) | 491 (18) |  |
| Native Hawaiian or Pacific Islander | 62 (2) | 2 (6) | 64 (2) |  | 58 (2) | 2 (3) | 60 (2) |  |
| White | 1,704 (50) | 9 (28) | 1,713 (50) |  | 1,338 (50) | 27 (38) | 1,365 (50) |  |
| Other | 200 (6) | 0 | 200 (6) |  | 101 (4) | 1 (1) | 102 (4) |  |
| Occupational risk level, count (%) |  |  |  | .786 |  |  |  | .464 |
| Low | 904 (26) | 7 (22) | 911 (26) |  | 738 (28) | 16 (23) | 754 (27) |  |
| Medium | 627 (18) | 7 (22) | 634 (18) |  | 477 (18) | 16 (23) | 493 (18) |  |
| High | 1,895 (55) | 18 (56) | 1,913 (55) |  | 1,468 (55) | 39 (55) | 1,507 (55) |  |
| Symptoms, count (%) |  |  |  |  |  |  |  |  |
| Fever | 391 (11) | 12 (38) | 403 (12) | < .001 | 245 (9) | 32 (45) | 277 (10) | < .001 |
| Cough | 562 (16) | 8 (25) | 570 (16) | .144 | 416 (16) | 25 (35) | 44 (16) | < .001 |
| Sore Throat | 645 (19) | 8 (25) | 653 (19) | .246 | 449 (17) | 21 (30) | 470 (17) | .006 |
| Runny Nose | 474 (14) | 8 (25) | 482 (14) | .067 | 370 (14) | 22 (31) | 392 (14) | < .001 |
| Loss of Smell | 67 (2) | 15 (47) | 82 (2) | < .001 | 38 (1) | 25 (35) | 63 (2) | < .001 |
| Loss of Taste | — | — | — | — | 42 (2) | 24 (34) | 66 (2) | < .001 |

Note. Group difference testing was performed with Mann-Whitney U tests for age and with Fisher's exact tests for categorical measures.

Nonparametric tests for group differences were performed for demographics and six symptoms of COVID-19. Significant differences between observed negative vs. positive cases at initial assessment were found for age (z = 2.396), race, fever, and loss of smell. At 8-week follow-up, significant differences were found for age (z = 4.718), race, and all symptoms (*p*’s < .05). Occupational risk did not contribute significantly to negative vs. positive group differences at either blood draw time point.

Among first responders’ initial blood draw, 12 antibody positive cases (214 negative) were identified, with an observed prevalence of 5.31% (exact binomial 95% CI = 2.77% - 9.09%). Accounting for test sensitivity and specificity, an adjusted prevalence of 5.75% (exact binomial 95% CI = 3.10% - 9.64%) was calculated, indicating 13 positive cases (213 negative) after adjustment. Significant differences were found for the symptoms of fever, cough, and loss of smell (*p*’s < .05). At their 8-week follow-up blood draw (n = 92), 4 antibody positive cases (88 negative) were identified, with an observed prevalence of 4.35% (exact binomial 95% CI = 1.20% - 10.76%) – an original 1 case remained antibody positive (11 did not return for the second blood draw) with an additional 3 new cases during an 8-weeks period (**Table 1b**). Adjusted prevalence was equal to observed prevalence. See **Table 3** for first responder sample characteristics and group differences.

**Table 3.** Sample Characteristics and Group Differences for First Responders at Baseline and 8-Week Follow-Up Assessments.

|  | Baseline |  |  |  | 8-Week Follow-Up |  |  |  |
| --- | --- | --- | --- | --- | --- | --- | --- | --- |
|  | Antibody Negative | Antibody Positive | Total | *p* | Antibody Negative | Antibody Positive | Total | *p* |
| Sample Size (%) | 214 (95) | 12 (5) | 226 (100) |  | 88 (96) | 4 (4) | 92 (100) |  |
| Age in yrs., *M* (*SD*) | 42.24 (8.63) | 38.33 (7.75) | 42.04 (8.61) | .206 | 41.91 (8.42) | 45.25 (2.22) | 42.05 (8.27) | .287 |
| Female, count (%) | 19 (9) | 1 (8) | 20 (9) | .713 | 12 (14) | 0 | 12 (13) | .566 |
| Race, count (%) |  |  |  | 1.000 |  |  |  | 1.000 |
| American Indian or Alaska Native | 0 | 0 | 0 |  | 0 | 0 | 0 |  |
| Asian | 12 (6) | 0 | 12 (5) |  | 4 (5) | 0 | 4 (4) |  |
| Black | 1 (0) | 0 | 1 (0) |  | 0 | 0 | 0 |  |
| Hispanic or Latino | 30 (14) | 1 (8) | 31 (14) |  | 7 (8) | 0 | 7 (8) |  |
| Native Hawaiian or Pacific Islander | 2 (1) | 0 | 2 (1) |  | 2 (2) | 0 | 2 (2) |  |
| White | 166 (78) | 11 (92) | 177 (78) |  | 75 (85) | 4 (100) | 79 (86) |  |
| Other | 3 (1) | 0 | 3 (1) |  | 0 | 0 | 0 |  |
| Symptoms, count (%) |  |  |  |  |  |  |  |  |
| Fever | 40 (19) | 6 (50) | 46 (20) | .018 | 15 (17) | 4 (100) | 19 (21) | .001 |
| Cough | 55 (26) | 8 (67) | 63 (28) | .005 | 22 (25) | 2 (50) | 24 (26) | .278 |
| Sore Throat | 49 (23) | 4 (33) | 53 (23) | .301 | 20 (23) | 1 (25) | 21 (23) | .652 |
| Runny Nose | 41 (19) | 5 (42) | 46 (20) | .072 | 22 (25) | 1 (25) | 23 (25) | .691 |
| Loss of Smell | 7 (3) | 6 (50) | 13 (6) | < .001 | 1 (1) | 0 | 1 (1) | .957 |
| Loss of Taste | — | — | — | — | 2 (2) | 0 | 2 (2) | .914 |

Note. Group difference testing was performed with Mann-Whitney U tests for age and with Fisher's exact tests for categorical measures.

Given our observed 8-week antibody persistence in HCWs, we also conducted an extrapolated prevalence calculation for the 8-week follow-up to include those with antibody positives at the first blood draw and who did not return for the second draw (see Table 1). For HCWs, adding these 4 cases (total positive n = 75) resulted in a prevalence of 2.72% (exact binomial 95% CI = 2.14% - 3.40%). Similarly adding 11 cases in the first responders (positive n = 15) resulted in a prevalence of 14.56% (exact binomial 95% CI = 8.39% - 22.88%). **Table 4** summarizes observed, adjusted, and extrapolated prevalence.

**Table 4.**
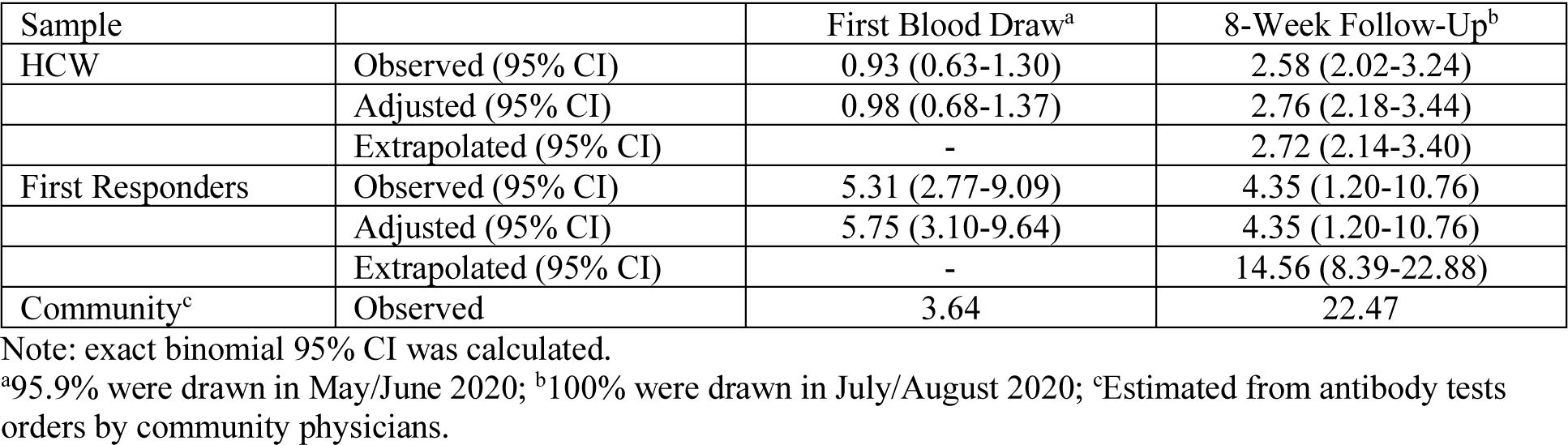
COVID-19 Prevalence Summary.

Among HCWs with previously PCR-confirmed diagnosis of COVID-19 (n = 75), 40 (53.3%) were antibody positive with 35 negative (46.7%) at 8-week follow-up. None of the negative 35 had a history of hospitalization or severe illness. While gender, race, and occupational risk did not significantly contribute to group differences between antibody negatives vs. positives, age and frequency of all symptoms were significantly different (*p*’s < .05), with positives significantly younger and presenting more symptoms than negatives. Among those with available PCR test date, time between PCR and antibody test ranged from 16 to 94 days (*M* = 41.33, *SD* = 23.27) for the negatives (n = 9) and from 12 to 151 days (*M* = 59.69, *SD* = 41.90) for the positives (n = 35), with no significant difference, *t*(42) = −1.26, *p* = .215. **Table 5** summarizes group differences.

**Table 5.**
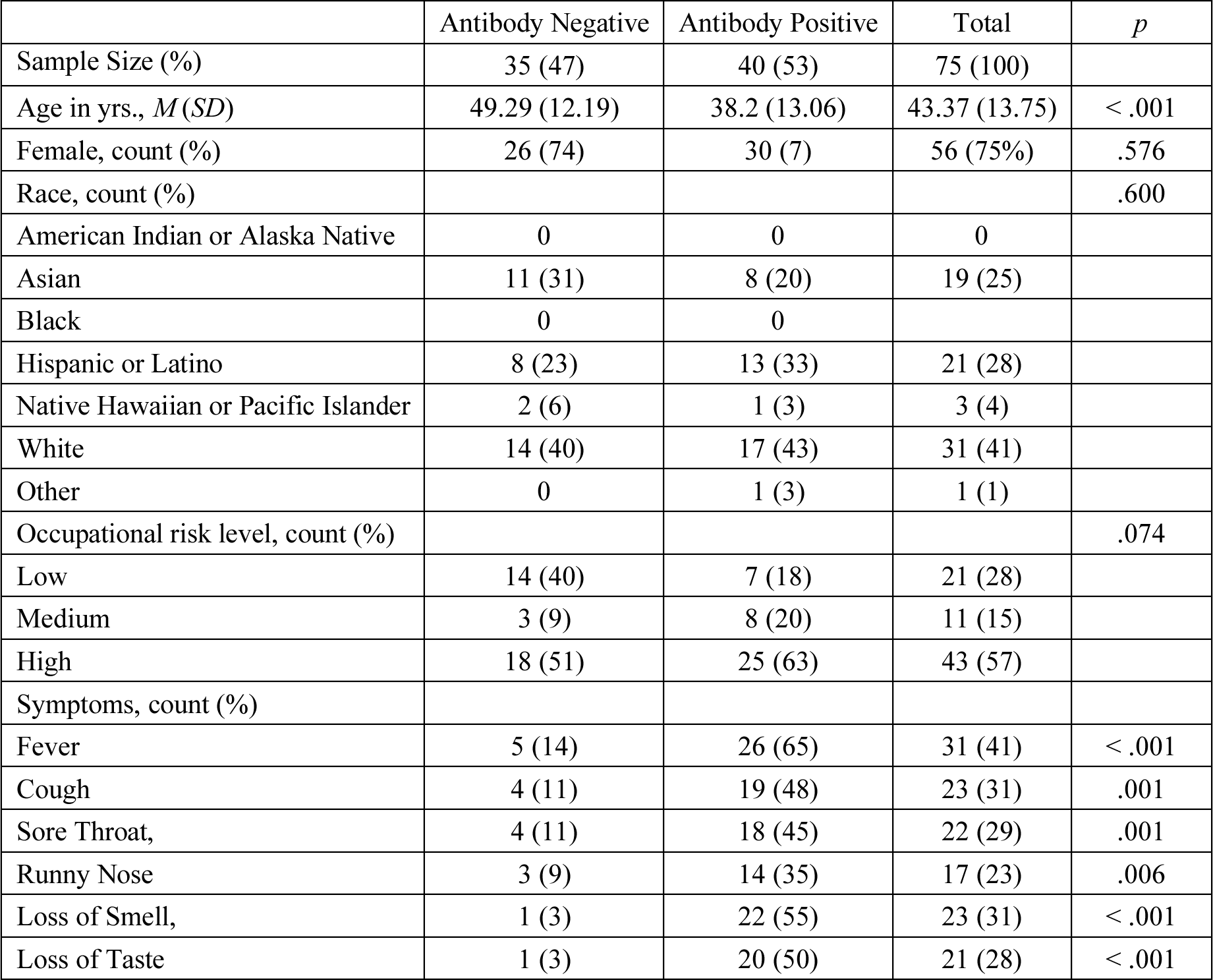
Sample Characteristics and Group Differences for 8-Week Follow-Up of HCWs with Prior PCR-confirmed COVID-19.

## DISCUSSION

Compared to other published studies, the present study found a considerably lower adjusted antibody prevalence (0.98%) on initial sampling (95.9% blood drawn in May and June) among HCWs as previously reported [19]. The community prevalence during this early period, when still under “shelter at home” orders under the Governor of California, was relatively low (3.64%) as reflected by those tested by physician order in our community, and this is reflected in adjusted prevalence among the first responders tested in this study (5.75%), although it is considerably higher than that of HCWs. While selection bias likely affected our estimate of community prevalence in serum drawn from physician orders, the antibody prevalence studies in local Southern California support our estimate [11,12]. The second period (100% blood drawn in July and August) reflected wider community transmission after partial state re-opening, as evidenced by a spike of hospitalization in Orange County, yet the higher adjusted prevalence rate in our HCWs (2.76%) was still well below of the considerably higher prevalence (22.47%), estimated from physician orders, in our community.

Several factors for the relatively low sero-prevalence in our HCWs may explain current findings. Upon reporting of the first COVID-19 patient in California (the third in the United States), our organization reacted immediately. We established an internal, weekly COVID-19 task force meeting and opened regular communication with the Orange County Healthcare Agency as well as the CDC to stay current with the rapidly changing guidelines from county, state, and federal agencies. The task force oversaw to date a rigorous approach to preparedness, including resource allocation (e.g., personal protective equipment, cohorted emergency room and hospital beds as well as ICU beds, dedicated staff) and hospital triage and process protocols, environmental cleansing and dietary rigor, rigorous visitation policies, all to amplify patients’ and workforces’ safety and infection prevention measures. Mandatory employee education and training on safety measures and prevention were implemented, heightening awareness among employees not only at work but more importantly outside of their work place. All those efforts may have contributed to this lower prevalence.

A relatively low regional estimated overall prevalence of infections in Orange County (total population of 3.2 million) also contributed to this low prevalence. This geographical effect can be seen in high antibody prevalence in HCWs in New York city, New York, USA, Wuhan, China, and Bergamo, Italy, where much higher community prevalence was reported. When our data were compared, using the economic re-opening in our county as a cut-off, between May/June vs. July/August, the low observed prevalence for both our HCWs (0.93% vs. 2.58%) and those tested by physician orders (3.6% vs. 22.5%) was reinforced. Incidentally, this trend was not observed for first responders, possibly due to smaller sample size and a large percentage of non-returning subjects at 8-week follow-up (although our expletory prevalence calculation did show this trend - 5.31% vs. 14.56%). Therefore, regional consideration must be given when considering antibody prevalence in HCWs.

Another possible explanation for lower susceptibility to infection among HCWs is the pre-existing presence of innate immunity [24] in HCWs acquired through T-cell mediated^25^ cross-reactivity to more common coronaviruse species [26-28]. This hypothesis postulates that greater exposure to such predecessors is experienced more commonly in hospital settings than the community at large. Recent studies document up to a 30% presence of such innate immunity in non-infected family members of those with confirmed infection. Prevalence of such pre-existing innate immunity in sampled blood donors prior to the epidemic has also been documented [6,29]. This phenomenon of innate immunity may also help explain the relatively low rate of infection susceptibility in younger children, given the common exposure to every day viral infections in pre-school and grade school [30-32].

Among HCWs with self-reported PCR-confirmed COVID-19, 46.7% were antibody negative (Table 5), which cannot be fully explained by antibody test sensitivity and specificity itself. Recent studies found a rapid decay of IgG antibody within the possible span of 2-3 months in patients with milder COVID-19 symptoms [24]. This may support our findings of the negative cases with significantly fewer symptoms compared to the positives. It should be noted that the loss of antibody positivity is not equivalent to loss of immunity [24]. This finding warrants further research in a larger cohort.

## CONCLUSIONS

Our findings suggest that the recommended in-patient personal protective equipment is effective in reducing the risk to HCWs and raising the confidence in those who need hospital care for urgent conditions to not delay seeking it. Also, the unexpected finding of lower rates of serologic conversion in our HCWs suggests the possibility that innate immunity may be greater among the HCWs, a hypothesis warranting further studies. Finally, the fact that all of our sero-positive HCWs have maintained antibody positivity for at least 8 weeks, with no reported re-infection, is encouraging, given the earlier reports of antibody evanescence [33,34].

## Data Availability

Data will be made available upon acceptance of the manuscript at the current submission.

## ACKNOWLEDGEMENT

We acknowledge our healthcare workers who have contributed to this study.

